# Changes in Fentanyl Distribution in California

**DOI:** 10.1101/2022.10.06.22280803

**Authors:** Miah V. Dugan, Ali H. Shah, Trinidy R. Anthony, Rafiat Famosa, Brian J. Piper

## Abstract

**Background:** Fentanyl is a synthetic opioid that is commonly given as a medication to alleviate pain. This drug can be administered through multiple routes, hence making it easy to exploit at high rates. Due to the flexibility in which it can be taken, it increases the ease of both access and use. The purpose of this study was to analyze trends in the distribution of fentanyl and its formulations across Medicaid enrollees in California and among the 3-digit registrant zip codes in California over the period of pre-pandemic (2018–2019) to the early stages of the COVID-19 pandemic (2020).

**Methods:** Using the Automated Reports and Consolidated Ordering System (ARCOS), the distribution of fentanyl across California was compiled from 2018 to 2020. Utilizing ARCOS, the number of individuals within the source population who lived in one of California’s many zip codes was observed. To analyze the fentanyl distribution trend, we used Google Sheets, GraphPad Prism (Version 9.3.0 [463]), and Microsoft 365 Excel. These were helpful to organize the Medicaid, ARCOS data, and as well as to create graphs. The Medicaid database was used to compile the number of fentanyl formulations prescribed from 2018 to 2020 across California.

**Results:** The analyses from both databases provided insight into the difference in fentanyl distribution in California from the years 2018 to 2020. After looking further into the many 3-digit registrant zip codes as well as Medicaid enrollees, it was found that there was a decrease in the distribution of fentanyl and its formulations. Additionally, it was found that the distribution of fentanyl as a medication by business activities also decreased from 2018 to 2020.

**Conclusion:** The results indicate that there was more fentanyl being distributed and prescribed before the pandemic (2018– 2019). On the other hand, when we considered the effects of the pandemic, during 2020, there was quite a drastic decrease in the amount of fentanyl being prescribed and distributed to those living in California and those enrolled in Medicaid.

## Introduction

Fentanyl and its analogs are lipid-soluble opioids, first synthesized 70 years ago in the 1950s (1). Fentanyl is an opioid agonist that acts in the u-opioid receptor while producing analgesia. This opioid is 50 to 100 times stronger than morphine. A minuscule dose, such as 100 micrograms, can produce pain relief equal to 10 mg of morphine. Its properties and pharmacokinetics differ widely from other drugs. It is mainly used as a sedative for post-surgical patients, epilepsy patients, and patients with renal failure. It is also used to treat chronic pain patients who have developed a tolerance for other drugs.

Fentanyl can be administered in many ways such as intravenously, intramuscularly, transdermally, intranasally, and intrathecally (2). From pre-COVID-19 (2018–2019) to the period of early COVID-19 (2020), there was a change in the illicit use of fentanyl and its derivatives as well as its distribution. There were also changes in the distribution and use of fentanyl, there has been a great increase in the prescription and manufacturing of illegal fentanyl (3). Due to these issues, the dependence on opioids has also increased. The overprescription of opioids has been one of the contributors to this crisis adding to many states limiting the prescription amounts of opioids, like fentanyl, restrictions in the prescription, and mandating the use of a program to monitor use (3).

Another study used was the urine drug test results before COVID-19 (November 2019 – March 2020) and during COVID-19 (March 2020 – July 2020). They found that there was a large correlation between fentanyl drug use that increased from 3.80% to 7.32% from before the pandemic to during the national emergency (4). Fentanyl is primarily responsible for overdose deaths across the United States, and this can be in part due to the prescription of opioids for pain. However, the pandemic has brought on an increase in opioid use (5). Using the ARCOS and Medicaid databases, we looked at changes in the fentanyl distribution in Medicaid recipients in California.

California is the state of choice to study primarily because of its significance in death rates in the state. Along with this, California is the largest state in the country by population. Having California as the state of choice gave us the ability to have a wide variety of data to discuss throughout our research. In the state of California, laws were implemented to decrease the rate of prescriptions of fentanyl. Law AB 1751 makes it more difficult to receive opioids in different states while law AB 2789 will require physicians to write electronic prescriptions (6).

The rate of overdoses due to synthetic opioids like fentanyl and fentanyl analogs increased over 16% from 2018 to 2019 and increased from 2020 to 2021 (7). There was a gap in understanding why there was a spike in fentanyl. Previous studies from 2021 reported those who use drugs containing fentanyl tend to smoke it more often. There was a change in the demand and supply chain for fentanyl since 2010, leading to a shift from 2018 to 2019 from injecting opioids to smoking fentanyl in California. This shift was due to the convenience of smoking the opioid rather than injecting it. Between 2018-2020, preliminary data from the CDC indicated a continuous rise in opioid morbidity (7).

In 2020, there was a staggering increase in the use of fentanyl, if this continues, it will surpass the highest point of use, which was in 2016. Although fentanyl prescription has declined over time, the death rate of this opioid continues to increase (8).

Another study in 2021, indicated that the rate of overdose in California increased by 49% between 2019 to 2020 (9). During this time, there were greater than projected overdose deaths in California amongst all racial and ethnic groups. In particular, the largest increase was seen in the Black community. This is likely related to systematic bias in the healthcare system regarding prescribed opioids and disparities in access to healthcare (10). This may be due to the lack of accessibility to healthcare resources and rehabilitation centers within these communities, thus leading to increased stress levels.

Other studies have reported that individuals who misuse fentanyl often experience a withdrawal effect when the opioid is unavailable. This could be a contributing factor to the increase in demand (11). Given that fentanyl is known for its potency and the harmful effects after its use, the drug continues to be known as a misused substance (3). This would allow us to explore the effect of current interventions used to combat this ongoing issue. The increase in the instances of fentanyl overdoses and fatalities is a direct result of illicit drug use. The rates continue to increase even though the number of prescriptions decreases because users are accessing opioids illegally from drug dealers that smuggle them from countries like China, Mexico, and India (12).

## Methods

### Database

We used fentanyl prescription data from the Medicaid database in California. The Medicaid database includes the outpatient drug utilization data for Medicaid enrollees in each state from the start of the drug rebate program (13). Other studies that used Medicaid showed a regional difference in the distribution of fentanyl from 2010 to 2019 (3). From the Automated Reports and Consolidated Ordering System (ARCOS), we used the fentanyl distribution by zip code within California for the years 2018 to 2020. Also, we used the retail purchases for fentanyl in grams by the business activity in California for the years 2018 to 2020 from the ARCOS database. The ARCOS database provides the distribution history of controlled drugs in the United States from the DEA (14). ARCOS provides a detailed overview of drug distribution by 3-digit zip code and business activity in the United States. Other studies that used ARCOS showed an overall decrease in distributed licit fentanyl from 2010 to 2019 and a decrease in fentanyl distribution by 17.9% from 2016 to 2017 (15,16). Our use of databases has helped find data collection specific to fentanyl and its analogs when determining the number of prescriptions given to those who are Medicaid patients; however, the use of outside sources has been beneficial to gaining an outside perspective (16). The purpose of this study was to examine how the distribution of fentanyl impacted Medicaid recipients in California from 2018 to 2020.

### Participants

The source population from the ARCOS database included individuals residing in one of the 57 zip codes in California. The target population included residents in one of the 3-digit zip codes of California who were distributed fentanyl. To further study changes in fentanyl distribution, the source population included Medicaid enrollees, and we focused on fentanyl users in the state of California. The target population consisted of Medicaid enrollees in California who were prescribed fentanyl. All individuals enrolled in Medicaid from 2018 to 2020 were included in this study. Persons that did not match our inclusion criteria for Medicaid and ARCOS were excluded from this analysis.

### Procedures

To determine the changes in fentanyl distribution in Medicaid users in California from 2018 to 2020, data was collected and sorted yearly from Medicaid. The number of prescriptions for each formulation was assessed through the state drug utilization Medicaid database from 2018 to 2020. Medicaid provides statistics for Medicaid enrollees and outpatient drug prescriptions in each state yearly. Any blank prescription values were replaced with zero.

To further analyze the distribution of fentanyl in California, data was collected from the ARCOS database from 2018 to 2020. The total number of prescriptions of fentanyl per 3-digit zip code in California was organized quarterly from 2018 to 2020. We assessed the total number of grams of fentanyl base distributed amongst pharmacies, hospitals, practitioners, teaching institutions, mid-level practitioners, and narcotic treatment programs.

### Statistics

We calculated the change in the percentage of fentanyl prescriptions from 2018 to 2020 (Table 1). To display the change in percentages of fentanyl prescriptions, we used a line graph and table to illustrate the data set collected from Medicaid. Along with the use of the Medicaid data, we also used data from the ARCOS database to see the amount of controlled use of the substance fentanyl in grams across the various zip codes in California. We performed a 95% confidence interval to evaluate if there was a significant difference between the means of the data, by determining if the p-value ≤0.05.

**Table 1.**
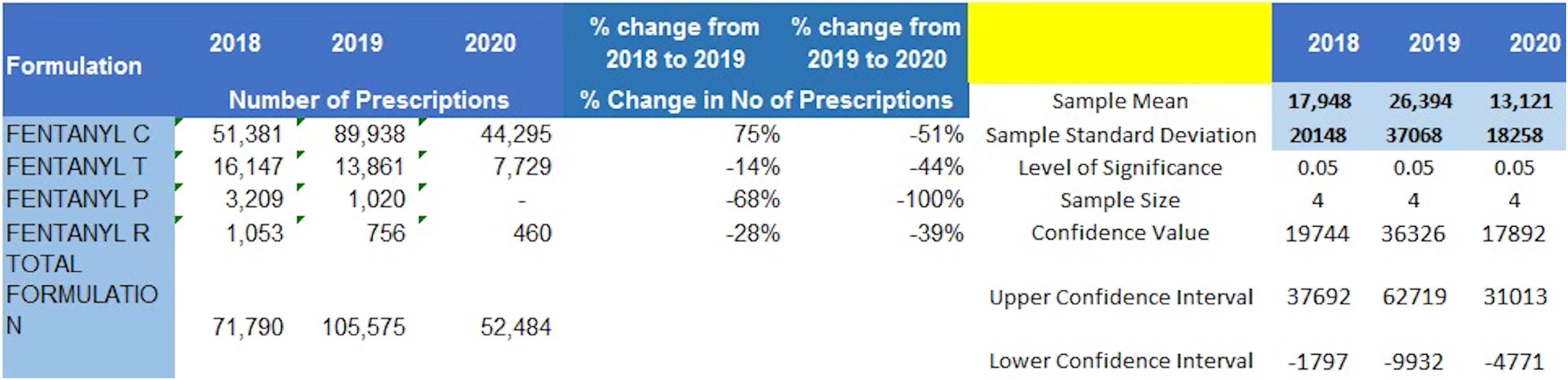
Changes in prescribed fentanyl prescriptions to Medicaid enrollees in California from 2018 to 2020.

The variables included in the analysis of Medicaid patients were the number of prescriptions prescribed, fentanyl formulations, number of quarters, and the years 2018, 2019, and 2020. For ARCOS, the variables used were the total grams of fentanyl distributed to California’s various 3-digit zip codes, total grams of fentanyl by its drug code to California, the quarterly drug distribution for California per 100,000 population, and the cumulative drug distribution to California per 100,000 population. Data were collected by each quarter (1–4), for each of our desired years (2018–2020) to evaluate the changes from the pre-COVID-19 pandemic to the early stage of the COVID-19 pandemic (Figure 2).

**Figure 1.**
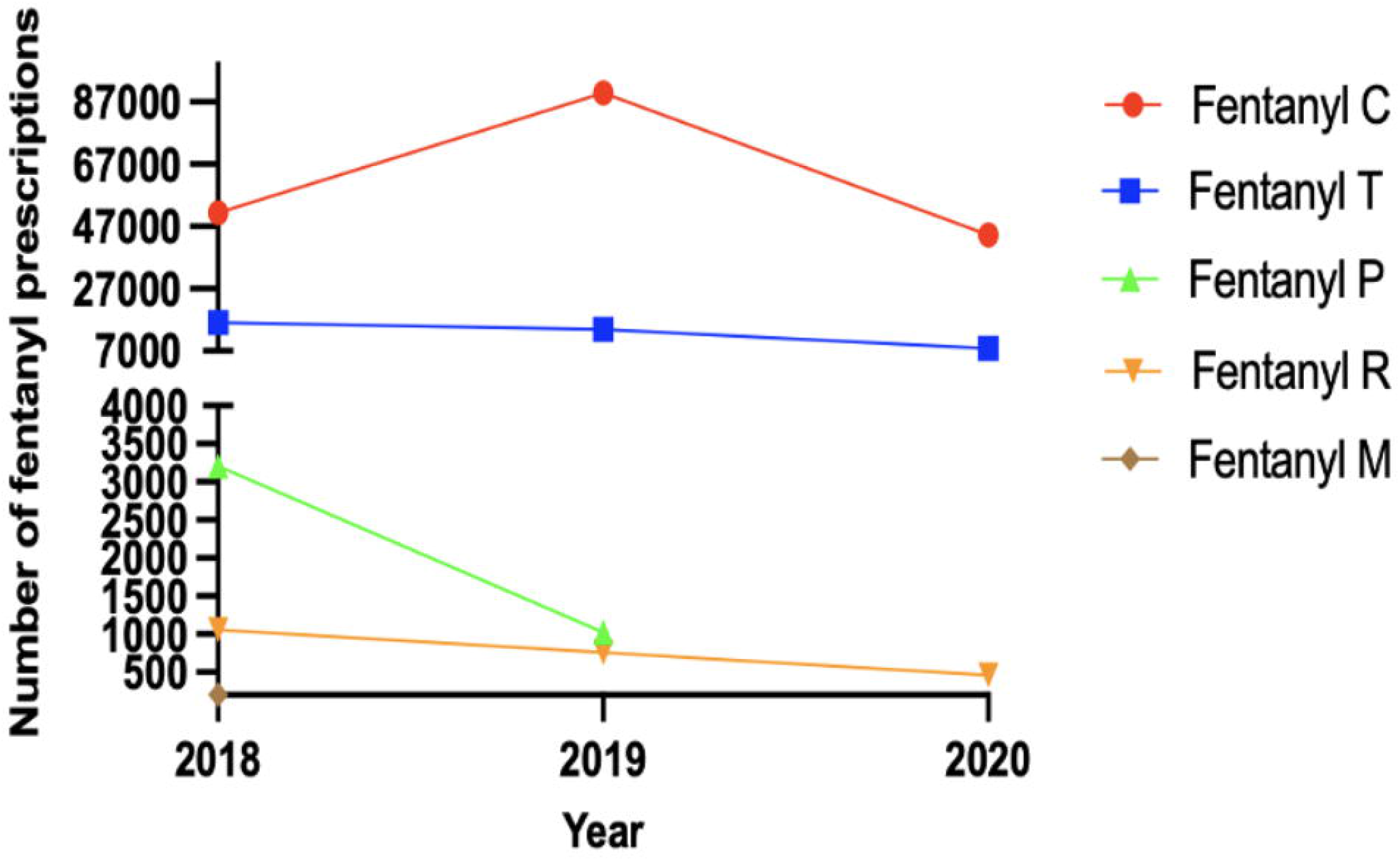
Number of fentanyl prescriptions prescribed for Medicaid patients in California from 2018-2020.

**Figure 2.**
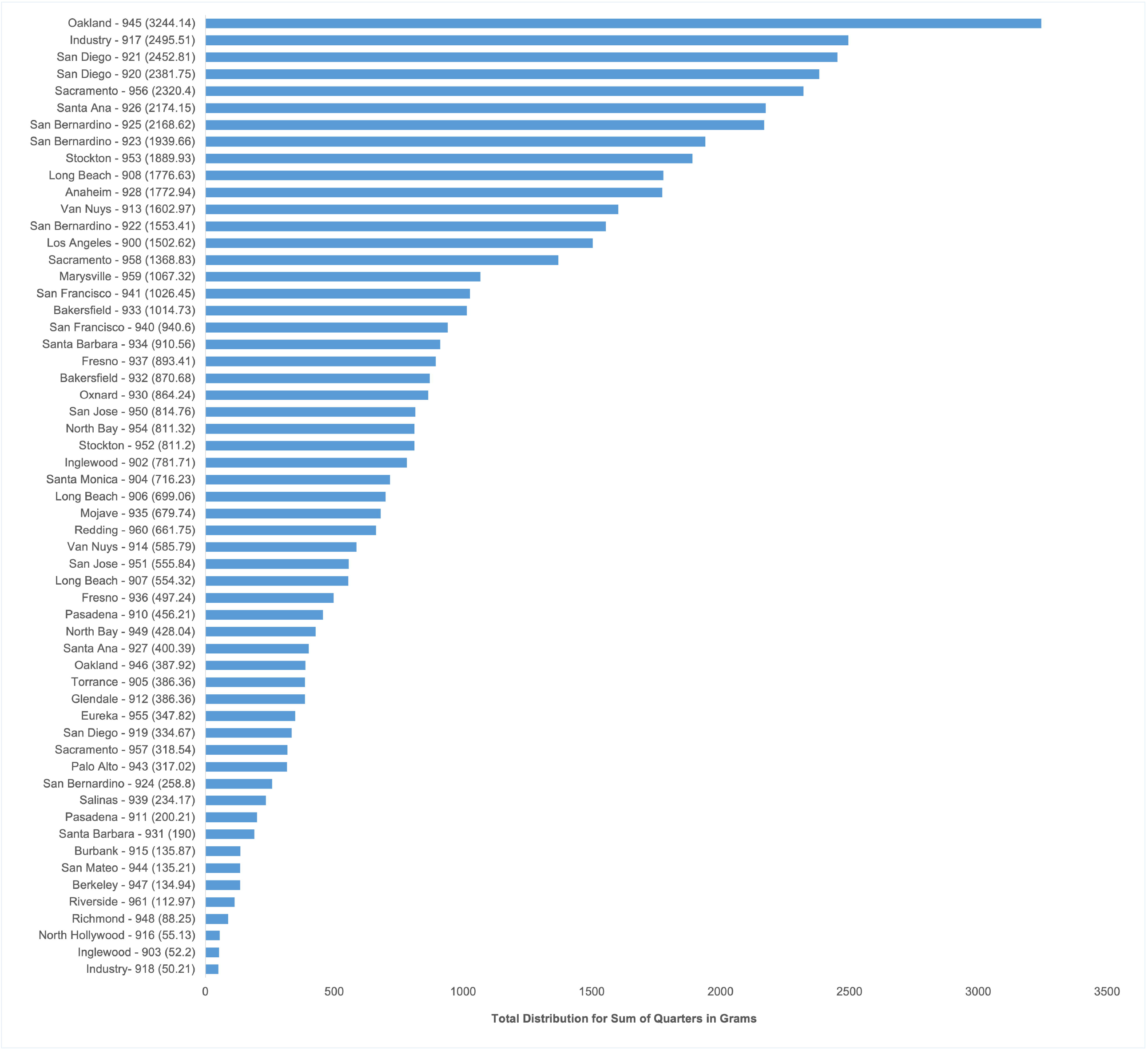
Total retail distribution of fentanyl drug base as a sum in California by zip code from 2018-2020 according to ARCOS.

To see the evident variations in the distribution of fentanyl, we created a visual representation of the sum of distributions for each 3-digit registrant zip code within California for each year (Figure 3). This was done to see how the distribution of fentanyl varied across each zip code within each year.

**Figure 3.**
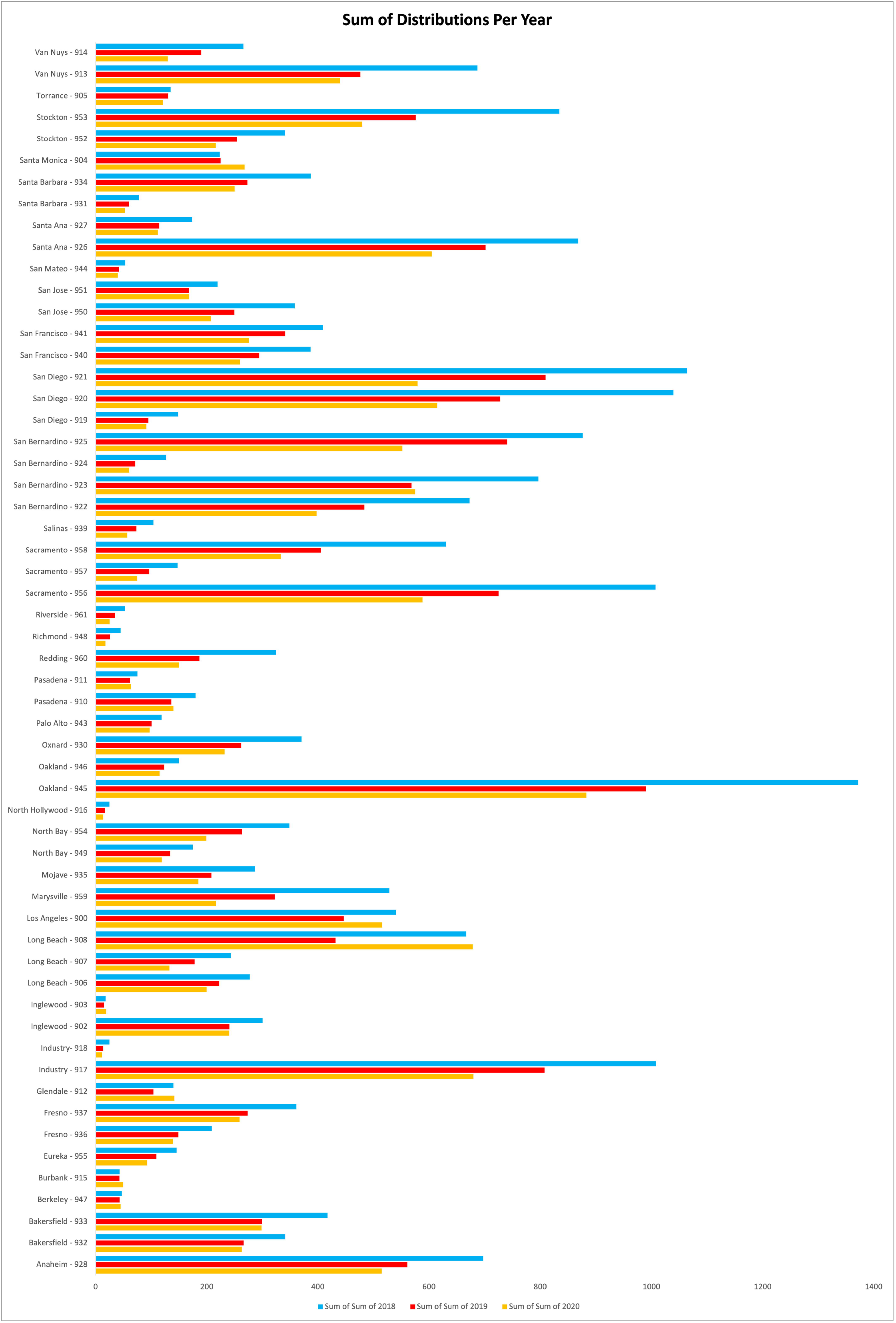
Retail distribution of fentanyl drug base as a sum in California by zip code for each year of 2018, 2019, and 2020 according to ARCOS.

GraphPad Prism (Version 9.3.0 [463]) and Microsoft 365 Excel were used for the statistical analysis and graphs. These programs were used to generate graphs and perform analyses. We formulated a heat map of the various zip codes of California to show how the corresponding zip codes for California correlate to the total amount of fentanyl distributed in grams (Figure 4). Business activity was measured to show the different amounts of fentanyl being distributed from various types of businesses.

**Figure 4.**
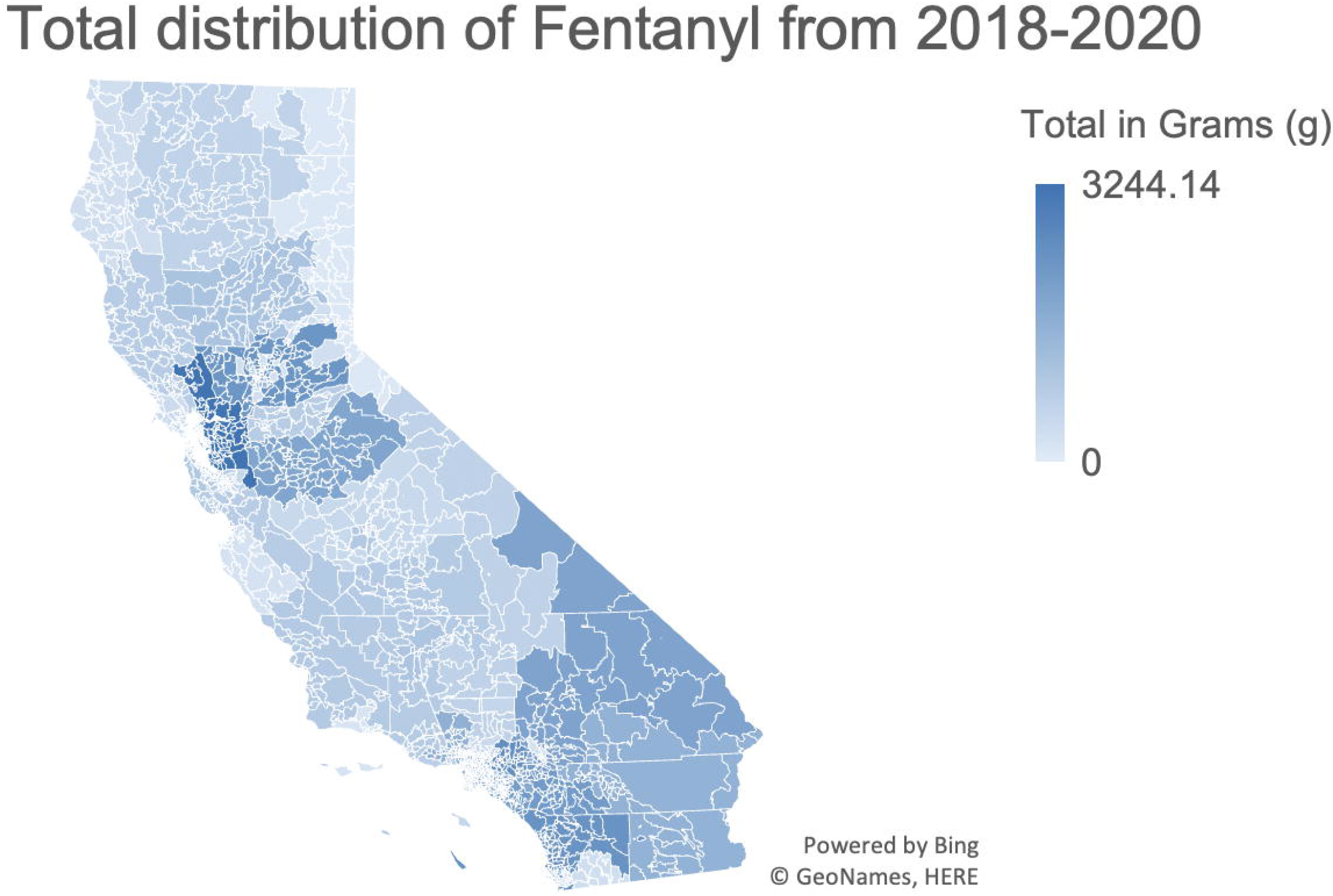
Heat map of total fentanyl distribution in California by zip code from 2018 to 2020 according to ARCOS.

## Results & Discussion

Table 1 demonstrates the change in the percentage of prescriptions given to those enrolled in Medicaid in California from 2018 to 2020. There is a clear decrease in the total number of prescriptions for each of the fentanyl formulations. The percentage difference in the number of prescribed formulations of fentanyl is also shown in Table 1. In addition, we incorporated the values for the 95% lower and higher confidence intervals to show the significance of our data. After analyzing the data, the decrease in fentanyl prescription could be due to the possibility that it is overused in various ways.

Figure 1 shows the decrease in the total number of fentanyl prescriptions for those enrolled in Medicaid for these years. Figure 2 demonstrates the total retail distribution of the drug base fentanyl as a sum by the 3-registrant zip codes within California. This shows the total amount of fentanyl that was distributed to each zip code during all three years (2018–2020). Figure 2 distinguishes zip codes that received a large amount of fentanyl compared to those that received much less.

Figure 3 shows the sum of fentanyl distribution for each of the three years and the evident transitions for fentanyl that was dispersed. This data analysis was also done for the state of California and by each 3-digit zip code. The heat map shown in Figure 4 displays the total amount of fentanyl allocated within the state of California by each of the zip codes for 2018 to 2020.

Figure 5 shows the business activity behind the distribution of fentanyl in California for the years 2018 to 2020. There is an apparent decrease in the distribution of fentanyl within California during these years in multiple business activities. This figure includes the amount of fentanyl distributed as a total in grams. The limitations to our study include not having the most recent data from 2021 due to factors such as the ongoing pandemic. This analysis does not account for those who are not enrolled in Medicaid and the data was in a de-identified form.

**Figure 5.**
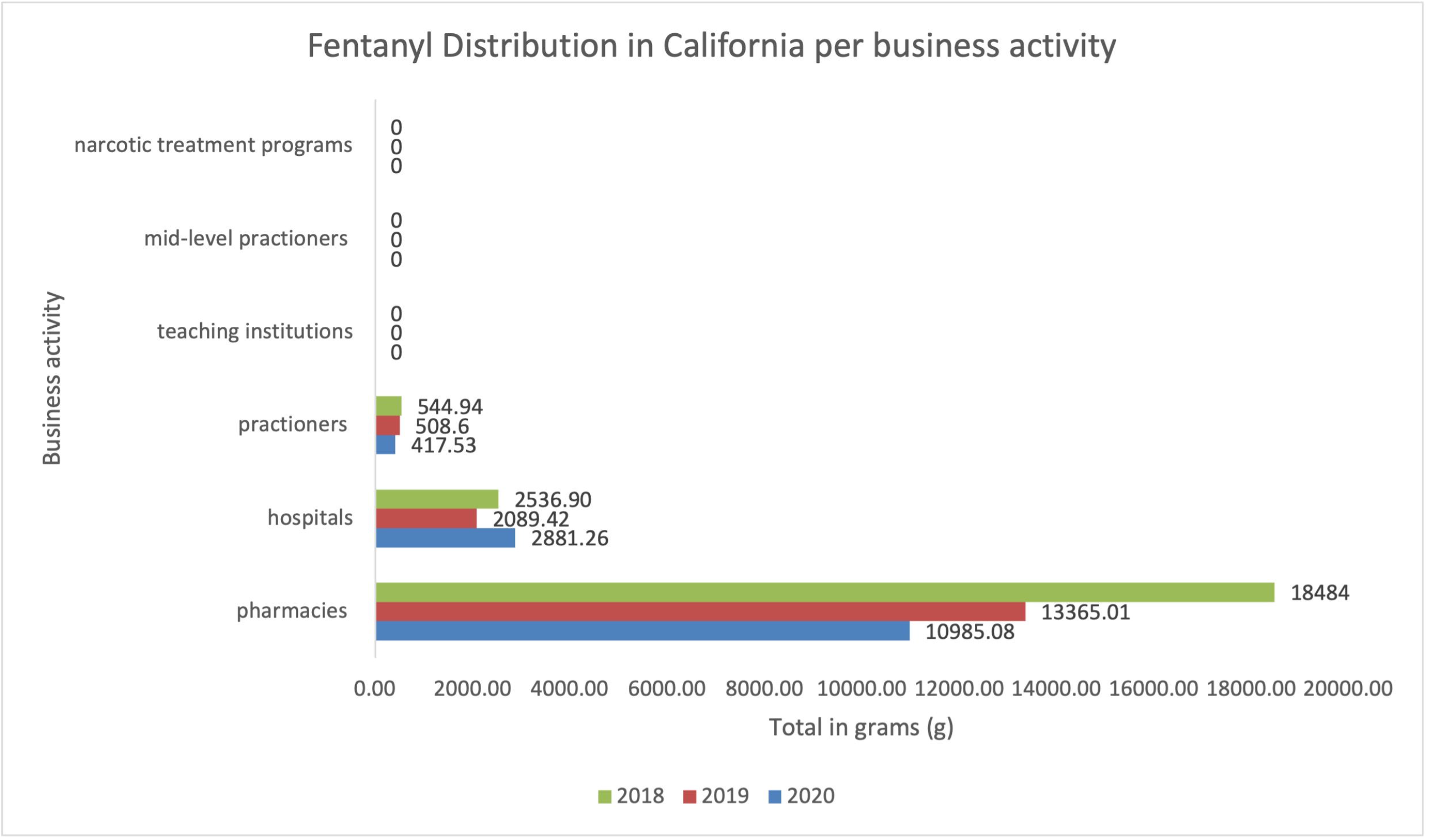
Total distribution of fentanyl in California by business activity.

Based on our findings, there was an increased distribution of fentanyl as well as the prescription of fentanyl and its formulations within the year 2018, which led to a decrease in the year 2019. Subsequently, once the pandemic began in 2020, there was a decrease in the same areas of analysis within California. Thus, the trend of decreasing distribution and prescription of fentanyl and its formulations occurred all within the year 2020.

## Conclusion

Our analysis concluded that there was a decrease in the distribution of fentanyl and the prescribing of fentanyl and its formulations. Even so, there was a spike in the prescription of fentanyl c formulation in 2019. Moreover, the geographical distinction is important in understanding the fentanyl distribution and prescription trends in California. The 3-digit zip code 945, Oakland, had the highest distribution of fentanyl for 2018, 2019, and 2020. The trend of decreasing distribution and prescription of fentanyl and its formulations can be seen as a possible correlation to the COVID-19 pandemic.

## Data Availability

All data produced in the present study are contained in the manuscript.

## Acknowledgments

The authors would like to thank Amy Houck for her extensive help in finding resources that fit our project’s main purpose.

## References

1. Stanley TH. Fentanyl. J Pain Symptom Manage. 2005;29(5 Suppl):S67–71. Available from: https://pubmed-ncbi-nlm-nih-gov.gcsom.idm.oclc.org/15907648/

2. Ramos-Matos C, Bistas K, Lopez-Ojeda W. Fentanyl [Internet]. Ncbi.nlm.nih.gov. 2022 [cited 27 May 2022]. Available from: https://www.ncbi.nlm.nih.gov/books/NBK459275/

3. Stemrich RA, Weber JV, McCall KL, Piper BJ. Pronounced declines in dispensed licit fentanyl, but not fentanyl derivatives. Res Social Adm Pharm. 2022 Jun;18(6):3046–3051. doi: 10.1016/j.sapharm.2021.08.001. Epub 2021 Aug 3.

4. Wainwright JJ, Mikre M, Whitley P, Dawson E, Huskey A, Lukowiak A, et al. Analysis of drug test results before and after the US Declaration of a National Emergency Concerning the COVID-19 outbreak. JAMA. 2020;324(16):1674–7. Available from: https://pubmed-ncbi-nlm-nih-gov.gcsom.idm.oclc.org/32945855/

5. Ciccarone D. The rise of illicit fentanyls, stimulants and the fourth wave of the opioid overdose crisis. Curr Opin Psychiatry. 2021;34(4):344–50. Available from: https://pubmed.ncbi.nlm.nih.gov/33965972/

6. Caiola S. Here are California’s new laws to address the state’s opioid crisis [Internet]. CapRadio. 2019 [cited 2022 Jul 18]. Available from: https://www.capradio.org/articles/2019/01/16/here-are-californias-new-laws-to-address-the-states-opioid-crisis/

7. Medicaid State Drug Utilization Data [Internet]. 1991 [cited 17 February 2022]. Available from: https://www.medicaid.gov/medicaid/prescription-drugs/state-drug-utilization-data/index.html.

8. ARCOS: Automation of Reports & Consolidated Orders System [Internet]. Department of Justice, Drug Enforcement Administration, 1980. 1980 [cited 17 February 2022]. Available from: https://www.deadiversion.usdoj.gov/arcos/index.html.

9. Collins LK, Pande LJ, Chung DY, Nichols SD, McCall KL, Piper BJ. Trends in the medical supply of fentanyl and fentanyl analogues: United States, 2006 to 2017. Prev Med. 2019;123:95–100. Available from: https://www.ncbi.nlm.nih.gov/pmc/articles/PMC8529416/

10. Young SD, Zhang Q, Zhou J, Pacula RL. Internet search and medicaid prescription drug data as predictors of opioid emergency department visits. NPJ Digit Med. 2021;4(1):21. Available from: https://pubmed-ncbi-nlm-nih-gov.gcsom.idm.oclc.org/33574500/

11. Kral AH, Lambdin BH, Browne EN, Wenger LD, Bluthenthal RN, Zibbell JE, et al. Transition from injecting opioids to smoking fentanyl in San Francisco, California. Drug Alcohol Depend. 2021;227:109003. Available from: https://pubmed-ncbi-nlm-nih-gov.gcsom.idm.oclc.org/34482046/

12. Manchikanti L, Vanaparthy R, Atluri S, Sachdeva H, Kaye AD, Hirsch JA. COVID-19 and the Opioid Epidemic: Two Public Health Emergencies That Intersect With Chronic Pain. Pain Ther. 2021;10(1):269–86. Available from: https://pubmed.ncbi.nlm.nih.gov/33718982/

13. Friedman J, Hansen H, Bluthenthal RN, Harawa N, Jordan A, Beletsky L. Growing racial/ethnic disparities in overdose mortality before and during the COVID-19 pandemic in California. Prev Med. 2021;153:106845. Available from: https://www.ncbi.nlm.nih.gov/pmc/articles/PMC8521065/

14. Kelley MA, Lucas J, Stewart E, Goldman D, Doctor JN. Opioid-related deaths before and after COVID-19 stay-at-home orders in Los Angeles County. Drug Alcohol Dependence. 2021;228:109028. Available from: https://pubmed.ncbi.nlm.nih.gov/34500239/

15. Blackwood CA, Cadet JL. COVID-19 Pandemic and fentanyl use disorder in African Americans. Front Neurosci. 2021;15:707386. Available from: https://pubmed-ncbi-nlm-nih-gov.gcsom.idm.oclc.org/34489626/

16. Intelligence Program DEA. Fentanyl flow to the United States [Internet]. DEA. 2020 [cited 2022Jul19]. Available from: https://www.dea.gov/documents/2020/2020-03/2020-03-06/fentanyl-flow-united-states

